# Diagnostic performance of CT and its key signs for COVID-19: A systematic review and meta-analysis

**DOI:** 10.1101/2020.05.24.20111773

**Authors:** Xiuting Wu, Yuhui Zhong, Wanyue Qin, Zhenxi Zhang, Kai Li

## Abstract

**Purpose:** To evaluate the diagnostic value of chest CT in 2019 novel coronavirus disease (COVID-19), using the reverse transcription polymerase chain reaction (RT-PCR) as a reference standard. At the same time, the imaging features of CT in confirmed COVID-19 patients would be summarized.

**Methods:** A comprehensive literature search of 5 electronic databases was performed. The pooled sensitivity, specificity, positive predictive value, and negative predictive value were calculated using the random-effects model and the summary receiver operating characteristic (SROC) curve. We also conducted a meta-analysis to estimate the pooled incidence of the chest CT imaging findings and the 95% confidence interval (95%CI). Meta-regression analysis was used to explore the source of heterogeneity.

**Results:** Overall, 25 articles comprising 4,857 patients were included. The pooled sensitivity of CT was 93% (95% CI, 89-96%) and specificity was 44% (95% CI, 27-62%). The area under the SROC curve was 0.94 (95% CI, 0.91-0.96). For the RT-PCR assay, the pooled sensitivity of the initial test and the missed diagnosis rate after the second-round test were 76% (95% CI: 59-89%; I^2^=96%) and 26% (95% CI: 14-39%; I^2^=45%), respectively. According to the subgroup analysis, the diagnostic sensitivity of CT in Hubei was higher than that in other regions. Besides, the most common patterns on CT imaging finding was ground glass opacities (GGO) 58% (95% CI: 49-70%), followed by air bronchogram 51% (95% CI: 31-70%). Lesions were inclined to distribute in peripheral 64% (95% CI: 49-78%), and the incidence of bilateral lung involvement was 69% (95% CI: 58-79%).

**Conclusions:** There were still several cases of missed diagnosis after multiple RT-PCR examinations. In high-prevalence areas, CT could be recommended as an auxiliary screening method for RT-PCR.

**Key points:**

1. Taking RT-PCR as the reference standard, the pooled sensitivity of CT was 93% (95% CI, 89-96%) and the specificity was 44% (95% CI, 27-62%). The area under the SROC curve was 0.94 (95% CI, 0.91-0.96).
2. For the RT-PCR assay, the pooled sensitivity of the initial test and the missed diagnosis rate after the second-round test were 76% (95% CI: 59-89%) and 26% (95% CI: 14-39%), respectively.
3. GGO was the key sign of the CT imaging, with an incidence of 58% (95% CI: 49-70%) in patients with SARS-CoV-2 infection. Pneumonia lesions were inclined to distribute in peripheral 64% (95% CI: 49-78%) and bilateral 69% (95% CI: 58-79%) lung lobes.

## 1. Introduction

Since December 2019, an unidentified pneumonia with fever, cough, and myalgia as clinical presentations emerged in some hospitals in Hubei, China [1]. Deep sequencing detection and analysis of respiratory samples revealed a species of novel coronavirus, known as acute respiratory syndrome coronavirus 2 (SARS-CoV-2) [2]. The disease, named COVID-19, can progress to acute respiratory distress syndrome in severe cases, that requires intensive care unit admission and oxygen therapy [3]. As of April 23, 2020, the Corona Virus has been spreading rapidly around the world, with more than 2475723 clinical-confirmed cases and 169151 confirmed death [4]. In the absence of a clear vaccine and specific antiviral treatments, early diagnosis and interrupting transmission became essential in dealing with the new emerging SARS-CoV-2.

The current recommendations for the diagnosis of COVID-19 are laboratory examinations such as nasopharyngeal and oropharyngeal swab tests. However, the problem of the false-negative RT-PCR assay for detecting SARS-CoV-2 was exposed in clinical work. It may be related to various aspects of nucleic acid detection experiments, such as sample collection, laboratory error, and so on. Until now, there are a variety of PCR kits available, with sensitivity ranging from 45% to 60%; thus, in the early stage of the disease, repeated tests may be required to obtain a definite diagnosis [5]. It is not easy to implement in global resource-limited settings. Since it mainly involved the respiratory system, a chest CT examination was strongly recommended for the preliminary assessment and follow-up of suspicious COVID-19 patients [6]. As a conventional imaging tool for diagnosing pneumonia, CT is relatively easier to operate and can quickly obtain diagnostic results. However, CT findings regarding these points showed variable diagnostic performance. For example, numerous cases had noteworthy CT findings despite a preliminary false-negative RT-PCR analysis results [7, 8]. Furthermore, several studies reported that CT had high sensitivity for COVID-19 [7, 9, 10], but a recent study including 121 cases found 56% of patients had a normal CT finding in the early stage of infection [11]. Hope et al. [12] described that CT did not add value to the diagnosis, while medical staff and CT scanners might also become infection vectors for other vulnerable patients who needed imaging.

Considering the inconsistency of the existing literature, we conducted a systematic review regarding the diagnostic performance of CT for COVID-19. At the same time, with the increasing global attention to COVID-19 outbreaks, a better understanding of chest CT imaging features is critical to ensuring patient management and treatment effectiveness. Therefore, we compiled the information to summarize the key signs of CT in patients with COVID-19.

## 2. Methods

This systematic review was conducted according to the guidelines of Preferred Reporting Items for Systematic Review and Meta-Analysis (PRISMA). The protocol was registered on PROSPERO (CRD42020179689) [13].

### 2.1 literature searches

Searches were conducted in five medical databases including three English databases (PubMed, Embase, and Web of Science) and two Chinese databases (China Biology Medicine disc, and China National Knowledge Infrastructure). The search terms used: “COVID-19”, “2019 novel coronavirus disease”, “COVID-19 pandemic”, “COVID-19 virus infection”, “coronavirus disease-19”, “2019 novel coronavirus infection”, “2019-nCoV infection”, “coronavirus disease 2019”, “2019-nCoV disease”, “COVID-19 virus disease”, and “CT”, “Computed Tomography”, “Imaging”, “radiological”. The searches were concluded by April 16, 2020, and three independent reviewers evaluated search results.

### 2.2 Eligibility criteria

The included articles were subject to the following criteria: (a) Publications were full-text original articles (b) The study took the RT-PCR assay as a reference standard and proposed the sensitivity and/or specificity of CT. (c) sufficient data available to the chest CT imaging characteristic of patients with SARS-CoV-2 infection. The exclusion criteria were as follows: (a) insufficient relevant result parameters and research data about chest CT. (b) reviews of the literature, case reports, or series. (c) literature with restrictions on the type of population or age. (c) study with a sample size less than 5. For studies with overlapping data, only data from the study with the most appropriate date or the largest sample size was included.

### 2.3. Data Extraction and quality evaluation

Two authors (Wu XT, Qin WY) used standardized data tables to extract the following data independently. (1) study characteristics: authors, journal, date (Month/Day), the region of studies, number of patients, study design (prospective or retrospective), and basic characteristics of the reference standard and index test. (2) diagnostic performance of RT-PCR and CT: When possible, we collect data including true negatives, true positives, false negatives, and false positives about CT. Furthermore, information about the sensitivity of the RT-PCR was collected. (3) patient characteristics: gender, mean or median age, epidemiological history, fever, cough, chest CT imaging features (GGO, consolidation, crazy paving pattern, airways abnormalities, air bronchogram, interlobular septal thickening, lymphadenopathy, pleural effusion, lesion location, and distribution).

An author (Zhong YH) extracted data on the quality of study and the risk of bias. We evaluate the quality of this meta-analysis using the tool of Quality Assessment of Diagnostic Accuracy Studies-2 (QUADAS-2) [14].

### 2.4 Statistical analysis

The sensitivity and/or specificity of CT in suspected COVID-19 patients were obtained from each study. We considered it as positive when pulmonary infiltration could be seen on CT: A patient that RT-PCR confirmed as COVID-19 was considered a true positive (TP) case, whereas one or more RT-PCR assay showed negative was considered a false positive (FP) case. No abnormality in chest CT was considered negative: A patient that RT-PCR confirmed as COVID-19 was considered a false negative (FN) case, whereas one or more PCR assay showed negative was considered a true negative (TN) case. Sensitivity was determined based on formula TP/(TP+FN) and specificity based on formula TN/(TN+FP). Incidences were pooled by the Freeman-Tukey Double arcsine transformation [15]. Pooled data and 95% CI were carried out by a random effect model. The overall diagnostic accuracy was graphically exhibited by the area under the SROC curves [16].

We used the I^2^ statistic and Cochran’s Q-test to assess heterogeneity between studies. Several covariates were as follows: (1)region of studies (Wuhan or not); (2) size of the study population (≥100 or < 100); (3) Studies mentioned the time interval between reference standard and CT (yes or not). Publication bias was estimated using the Deeks funnel plot asymmetry test. All data analysis was conducted using the Stata 15.1 and “meta” package in R software version 3.6.3.

## 3. Results

### 3.1 Study Selection and characteristics

The process of study selection is described in Figure 1. The overall computer study search of the 5 databases yielded 1518 potentially relevant records. After removing duplicates and irrelevancies, there was still 63 literature, of which 723 were excluded base on titles or abstracts. At last, we read the full text of the remaining 63 documents carefully and excluded 38 studies. Therefore, a total of 25 papers were selected in this study (Table 1) [7, 9-11, 17-37].

**Figure 1:**
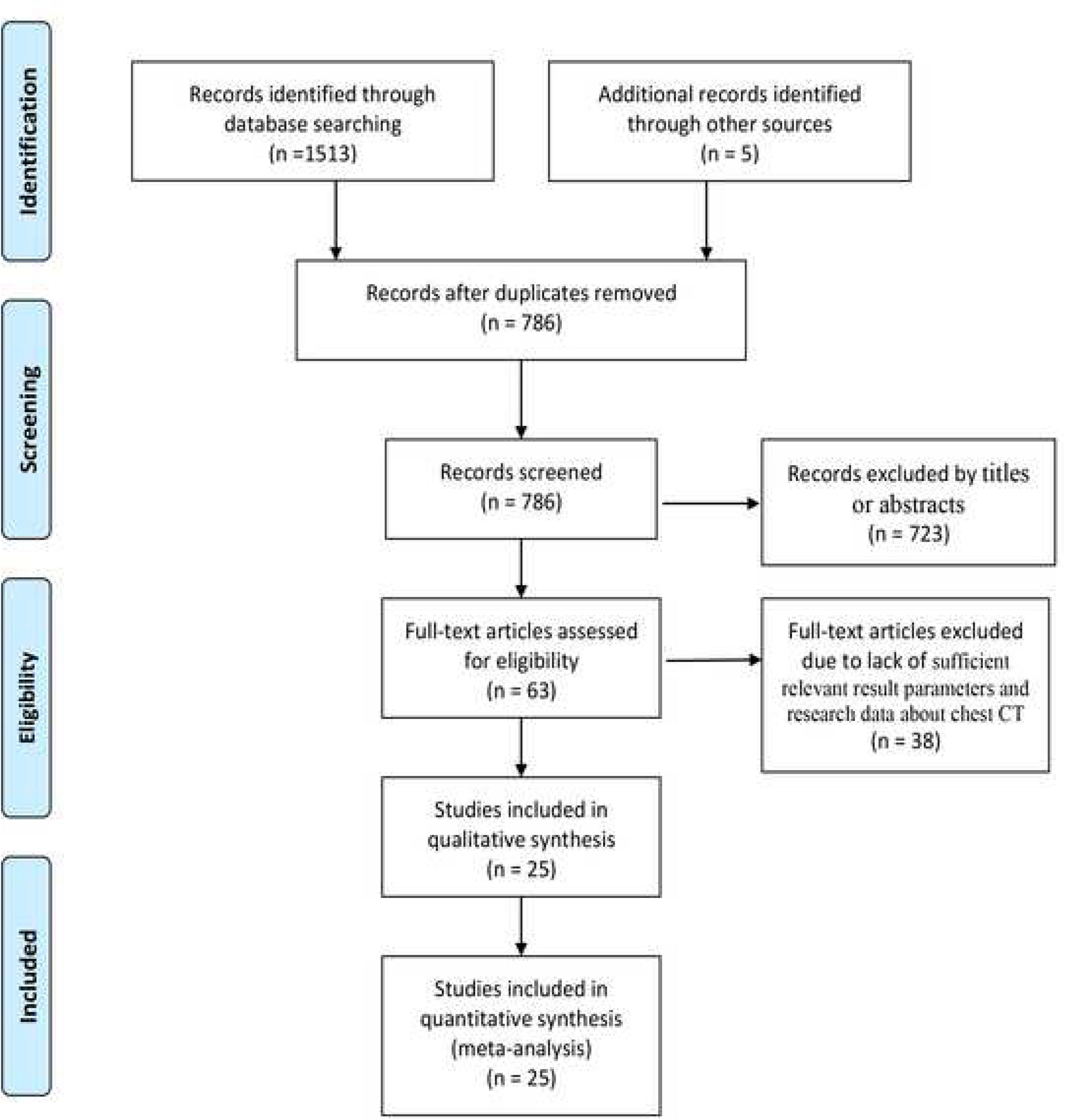
Flow diagram for study selection.

**Table 1:**
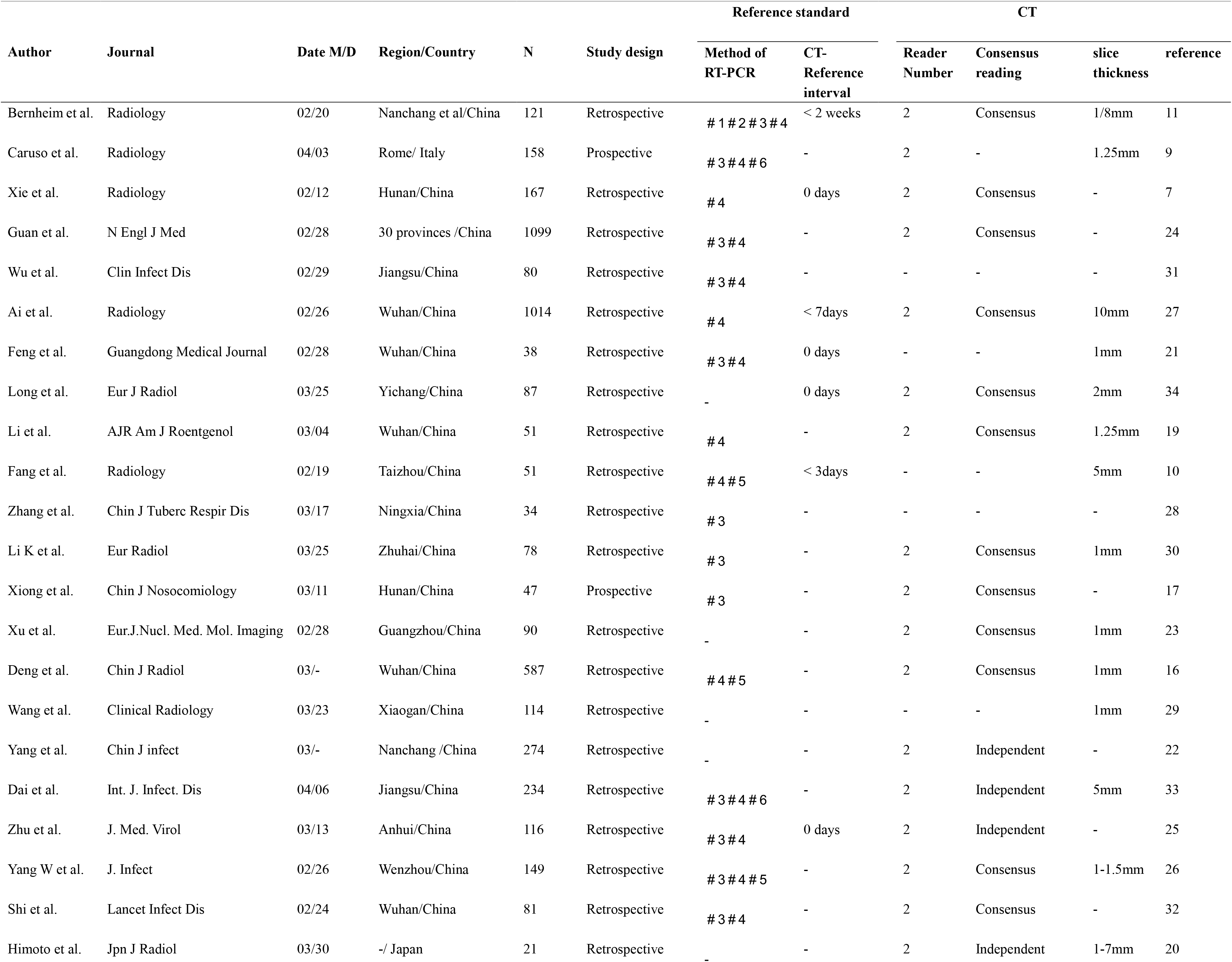
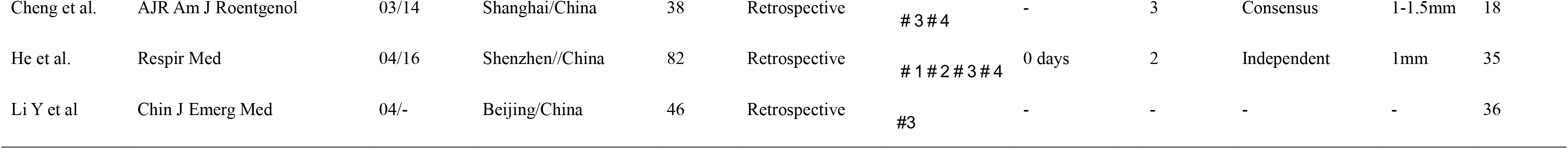
The characteristics of the literature included in meta-analysis. M/D=Month/Day, RT-PCR= reverse transcriptase polymerase chain reaction. Sample collection: #1=bronchoalveolar lavage; #2= endotracheal aspirate; #3= nasopharyngeal swab; #4= oropharyngeal swab; # 5= sputum samples; # 6= specimens from blood.

| Author | Journal | Date M/D | Region/Country | N | Study design | Reference standard |  | CT |  |  |  |
| --- | --- | --- | --- | --- | --- | --- | --- | --- | --- | --- | --- |
|  |  |  |  |  |  | Method of RT-PCR | CT-Reference interval | Reader Number | Consensus reading | slice thickness | reference |
| Bernheim et al. | Radiology | 02/20 | Nanchang et al/China | 121 | Retrospective | # 1 # 2 # 3 # 4 | < 2 weeks | 2 | Consensus | 1/8mm | 11 |
| Caruso et al. | Radiology | 04/03 | Rome/ Italy | 158 | Prospective | # 3 # 4 # 6 | - | 2 | - | 1.25mm | 9 |
| Xie et al. | Radiology | 02/12 | Hunan/China | 167 | Retrospective | # 4 | 0 days | 2 | Consensus | - | 7 |
| Guan et al. | N Engl J Med | 02/28 | 30 provinces /China | 1099 | Retrospective | # 3 # 4 | - | 2 | Consensus | - | 24 |
| Wu et al. | Clin Infect Dis | 02/29 | Jiangsu/China | 80 | Retrospective | # 3 # 4 | - | - | - | - | 31 |
| Ai et al. | Radiology | 02/26 | Wuhan/China | 1014 | Retrospective | # 4 | < 7days | 2 | Consensus | 10mm | 27 |
| Feng et al. | Guangdong Medical Journal | 02/28 | Wuhan/China | 38 | Retrospective | # 3 # 4 | 0 days | - | - | 1mm | 21 |
| Long et al. | Eur J Radiol | 03/25 | Yichang/China | 87 | Retrospective | - | 0 days | 2 | Consensus | 2mm | 34 |
| Li et al. | AJR Am J Roentgenol | 03/04 | Wuhan/China | 51 | Retrospective | # 4 | - | 2 | Consensus | 1.25mm | 19 |
| Fang et al. | Radiology | 02/19 | Taizhou/China | 51 | Retrospective | # 4 # 5 | < 3days | - | - | 5mm | 10 |
| Zhang et al. | Chin J Tuberc Respir Dis | 03/17 | Ningxia/China | 34 | Retrospective | # 3 | - | - | - | - | 28 |
| Li K et al. | Eur Radiol | 03/25 | Zhuhai/China | 78 | Retrospective | # 3 | - | 2 | Consensus | 1mm | 30 |
| Xiong et al. | Chin J Nosocomiology | 03/11 | Hunan/China | 47 | Prospective | # 3 | - | 2 | Consensus | - | 17 |
| Xu et al. | Eur.J.Nucl. Med. Mol. Imaging | 02/28 | Guangzhou/China | 90 | Retrospective | - | - | 2 | Consensus | 1mm | 23 |
| Deng et al. | Chin J Radiol | 03/- | Wuhan/China | 587 | Retrospective | # 4 # 5 | - | 2 | Consensus | 1mm | 16 |
| Wang et al. | Clinical Radiology | 03/23 | Xiaogan/China | 114 | Retrospective | - | - | - | - | 1mm | 29 |
| Yang et al. | Chin J infect | 03/- | Nanchang /China | 274 | Retrospective | - | - | 2 | Independent | - | 22 |
| Dai et al. | Int. J. Infect. Dis | 04/06 | Jiangsu/China | 234 | Retrospective | # 3 # 4 # 6 | - | 2 | Independent | 5mm | 33 |
| Zhu et al. | J. Med. Virol | 03/13 | Anhui/China | 116 | Retrospective | # 3 # 4 | 0 days | 2 | Independent | - | 25 |
| Yang W et al. | J. Infect | 02/26 | Wenzhou/China | 149 | Retrospective | # 3 # 4 # 5 | - | 2 | Consensus | 1-1.5mm | 26 |
| Shi et al. | Lancet Infect Dis | 02/24 | Wuhan/China | 81 | Retrospective | # 3 # 4 | - | 2 | Consensus | - | 32 |
| Himoto et al. | Jpn J Radiol | 03/30 | -/ Japan | 21 | Retrospective | - | - | 2 | Independent | 1-7mm | 20 |
| Cheng et al. | AJR Am J Roentgenol | 03/14 | Shanghai/China | 38 | Retrospective | # 3 # 4 | - | 3 | Consensus | 1-1.5mm | 18 |
| He et al. | Respir Med | 04/16 | Shenzhen//China | 82 | Retrospective | # 1 # 2 # 3 # 4 | 0 days | 2 | Independent | 1mm | 35 |
| Li Y et al | Chin J Emerg Med | 04/- | Beijing/China | 46 | Retrospective | #3 | - | - | - | - | 36 |

Table 1-2 list the characteristics of the included studies. This review of 25 studies included 4,857 patients, published from February 12, 2020, to April 16, 2020, most of them from China and two from Italy and Japan [9, 21]. Except for two studies [9, 18], the designs were all retrospective studies. Taking RT-PCR as the standard, 25 studies reported the diagnostic sensitivity of CT, and 11 studies reported the diagnostic specificity of CT [5, 9, 17-19, 21, 23, 26, 35-37]. Furthermore, there are 10 studies reported the diagnostic sensitivity of the initial PCR assay [7, 10, 11, 18, 22, 29, 32, 34-36], while 5 of them mentioned the rate of missed diagnosis after the second tests [10, 22, 29, 32, 35]. We analyzed a total of 33 variables for the meta-analyses (Table 3).

**Table 2:** Diagnostic performance of chest CT for COVID-19 using the RT-PCR as the reference standard. TP=true positive, TN=true negative, FP=false positive, FN=false negative, PPV= positive predictive value, NPV= negative predictive value.

| Author | N | Chest CT |  |  |  |  |  |  |  |  | Positive result of RT-PCR |  |  |  | reference |
| --- | --- | --- | --- | --- | --- | --- | --- | --- | --- | --- | --- | --- | --- | --- | --- |
|  |  | TP | TN | FP | FN | Sensitivity | Specificity | PPV | NPV | Accuracy | The first time | The Second time | The third or more times | Eventually positive result |  |
| Bernheim et al. | 121 | 94 | - | - | 27 | 0.78 | - | - | - | - | 90 | - | - | 121 | 11 |
| Caruso et al. | 158 | 60 | 54 | 42 | 2 | 0.97 | 0.56 | 0.59 | 0.96 | 0.72 | - | - | - | 62 | 9 |
| Xie et al. | 167 | 160 | - | - | 7 | 0.96 | - | - | - | - | 162 | - | - | 167 | 7 |
| Guan et al. | 1099 | 840 | - | - | 135 | 0.86 | - | - | - | - | - | - | - | 1099 | 24 |
| Wu et al. | 80 | 55 | - | - | 25 | 0.69 | - | - | - | - | 41 | 30 | 9 | 80 | 31 |
| Ai et al. | 1014 | 580 | 105 | 308 | 21 | 0.97 | 0.25 | 0.65 | 0.83 | 0.68 | - | - | - | 601 | 27 |
| Feng et al. | 38 | 37 | - | - | 1 | 0.97 | - | - | - | - | 9 | 25 | 4 | 38 | 21 |
| Long et al. | 87 | 35 | 0 | 51 | 1 | 0.97 | 0.00 | 0.41 | 0.00 | 0.40 | 30 | 3 | 3 | 36 | 34 |
| Li et al. | 51 | 49 | - | - | 2 | 0.96 | - | - | - | - | - | - | - | 51 | 19 |
| Fang et al. | 51 | 50 | - | - | 1 | 0.98 | - | - | - | - | 36 | 12 | 3 | 51 | 10 |
| Zhang et al. | 34 | 29 | - | - | 5 | 0.85 | - | - | - | - | 22 | 6 | 6 | 34 | 28 |
| Li K et al. | 78 | 56 | - | - | 22 | 0.72 | - | - | - | - | - | - | - | 78 | 30 |
| Xiong et al. | 47 | 19 | 19 | 8 | 1 | 0.95 | 0.70 | 0.70 | 0.95 | 0.81 | 14 | - | - | 20 | 17 |
| Xu et al. | 90 | 69 | - | - | 21 | 0.77 | - | - | - | - | - | - | - | 90 | 23 |
| Deng et al. | 587 | 423 | 83 | 71 | 10 | 0.98 | 0.54 | 0.86 | 0.89 | 0.86 | - | - | - | 433 | 16 |
| Wang et al. | 114 | 110 | - | - | 4 | 0.96 | - | - | - | - | - | - | - | 114 | 29 |
| Yang et al. | 274 | 48 | 151 | 70 | 5 | 0.91 | 0.68 | 0.41 | 0.97 | 0.73 | - | - | - | 53 | 22 |
| Dai et al. | 234 | 219 | - | - | 15 | 0.94 | - | - | - | - | 228 | - | - | 234 | 33 |
| Zhu et al. | 116 | 30 | 28 | 56 | 2 | 0.94 | 0.33 | 0.31 | 0.93 | 0.50 | - | - | - | 32 | 25 |
| Yang W et al. | 149 | 132 | - | - | 17 | 0.89 | - | - | - | - | - | - | - | 149 | 26 |
| Shi et al. | 81 | 81 | - | - | 0 | 1.00 | - | - | - | - | - | - | - | 81 | 32 |
| Himoto et al. | 21 | 6 | 3 | 12 | 0 | 1.00 | 0.20 | 0.33 | 1.00 | 0.43 | - | - | - | 6 | 20 |
| Cheng et al. | 38 | 11 | 5 | 22 | 0 | 1.00 | 0.19 | 0.33 | 1.00 | 0.42 | - | - | - | 11 | 18 |
| He et al. | 82 | 26 | 46 | 2 | 8 | 0.76 | 0.96 | 0.93 | 0.85 | 0.88 | 27 | - | - | 34 | 35 |
| Li Y et al | 46 | 9 | 21 | 16 | 0 | 1.00 | 0.57 | 0.36 | 1.00 | 0.65 | - | - | - | 9 | 36 |

**Table 3:** Meta-analysis outcomes (random-effects model). 95% CI = 95% confidence interval.

| Variable | Pooled<br>Proportion | 95% CI | p | I<sup>2</sup><br>(%) | t<sup>2</sup> |
| --- | --- | --- | --- | --- | --- |
| Diagnostic performance of CT |  |  |  |  |  |
| Sensitivity of CT | 0.93 | 0.89-0.96 | P<0.01 | 89 | 0.0164 |
| Specificity of CT | 0.44 | 0.27-0.62 | P<0.01 | 97 | 0.0808 |
| PPV | 0.55 | 0.41-0.68 | P<0.01 | 96 | 0.0450 |
| NPV | 0.97 | 0.91-1.00 | P<0.01 | 65 | 0.0099 |
| Diagnostic performance of RT-PCR |  |  |  |  |  |
| Sensitivity of initial RT-PCR assay | 0.76 | 0.59-0.89 | P<0.01 | 96 | 0.0762 |
| Missed diagnosis rate after<br>the second-round RT-PCR assay | 0.26 | 0.14-0.39 | P=0.12 | 45 | 0.0106 |
| Imaging features of chest CT |  |  |  |  |  |
| GGO | 0.58 | 0.45-0.70 | P<0.01 | 94 | 0.0401 |
| Air bronchogram | 0.51 | 0.31-0.70 | P<0.01 | 96 | 0.0720 |
| Interlobular septal thickening | 0.32 | 0.17-0.49 | P<0.01 | 95 | 0.0418 |
| Airways abnormalities | 0.31 | 0.07-0.61 | P<0.01 | 98 | 0.1724 |
| Linear opacities | 0.29 | 0.03-0.66 | P<0.01 | 97 | 0.1077 |
| Consolidation | 0.20 | 0.07-0.37 | P<0.01 | 94 | 0.0660 |
| Crazy-paving pattern | 0.17 | 0.04-0.36 | P<0.01 | 95 | 0.0714 |
| Nodules | 0.05 | 0.01-0.12 | P<0.01 | 88 | 0.0274 |
| Pleural effusion | 0.04 | 0.02-0.06 | P<0.01 | 63 | 0.0054 |
| Lymphadenopathy | 0.04 | 0.00-0.10 | P<0.01 | 92 | 0.0375 |
| Bilateral lung distribution | 0.69 | 0.58-0.79 | P<0.01 | 94 | 0.0291 |
| Peripheral distribution | 0.64 | 0.49-0.78 | P<0.01 | 94 | 0.0467 |
| 1 lobe involved | 0.09 | 0.06-0.12 | P=0.24 | 25 | 0.0009 |
| 2 lobes involved | 0.06 | 0.03-0.09 | P=0.26 | 25 | 0.0012 |
| 3 lobes involved | 0.08 | 0.05-0.12 | P=0.28 | 21 | 0.0010 |
| 4 lobes involved | 0.11 | 0.08-0.15 | P=0.42 | 0 | 0.0000 |
| 5 lobes involved | 0.48 | 0.30-0.66 | P<0.01 | 92 | 0.0432 |
| More than 2 lobes involved | 0.61 | 0.48-0.74 | P<0.01 | 90 | 0.0212 |
| Right upper lobe | 0.38 | 0.29-0.49 | P<0.01 | 82 | 0.0110 |
| Right middle lobe | 0.35 | 0.28-0.43 | P<0.01 | 70 | 0.0055 |
| Right lower lobe | 0.51 | 0.35-0.67 | P<0.01 | 93 | 0.0305 |
| Left upper lobe | 0.41 | 0.30-0.54 | P<0.01 | 88 | 0.0169 |
| Left lower lobe | 0.49 | 0.34-0.64 | P<0.01 | 92 | 0.0283 |
| Clinical characteristics |  |  |  |  |  |
| Gender(male) | 0.57 | 0.52-0.62 | P<0.01 | 74 | 0.0067 |
| Epidemiological history | 0.81 | 0.72-0.89 | P<0.01 | 91 | 0.0318 |
| Fever | 0.79 | 0.66-0.90 | P<0.01 | 97 | 0.0696 |
| Cough | 0.58 | 0.48-0.68 | P<0.01 | 93 | 0.0295 |

### 3.2 Quality assessment and Publication bias

In general, according to QUADAS-2, the 25 studies in this meta-analysis showed moderate methodological quality (Figure 2). Since most of the literature we included were retrospective studies, there was a high risk of bias for patient selection in 8 (32%) studies. The index tests of 14 studies showed low risk, while the rest of the studies did not know the risk of bias due to lack of information. The blind interpretation of index tests and reference standards in 12 studies is unclear. In several studies, the lack of description of reference standards was not considered to raise concerns about the applicability of the results.

**Figure 2:**
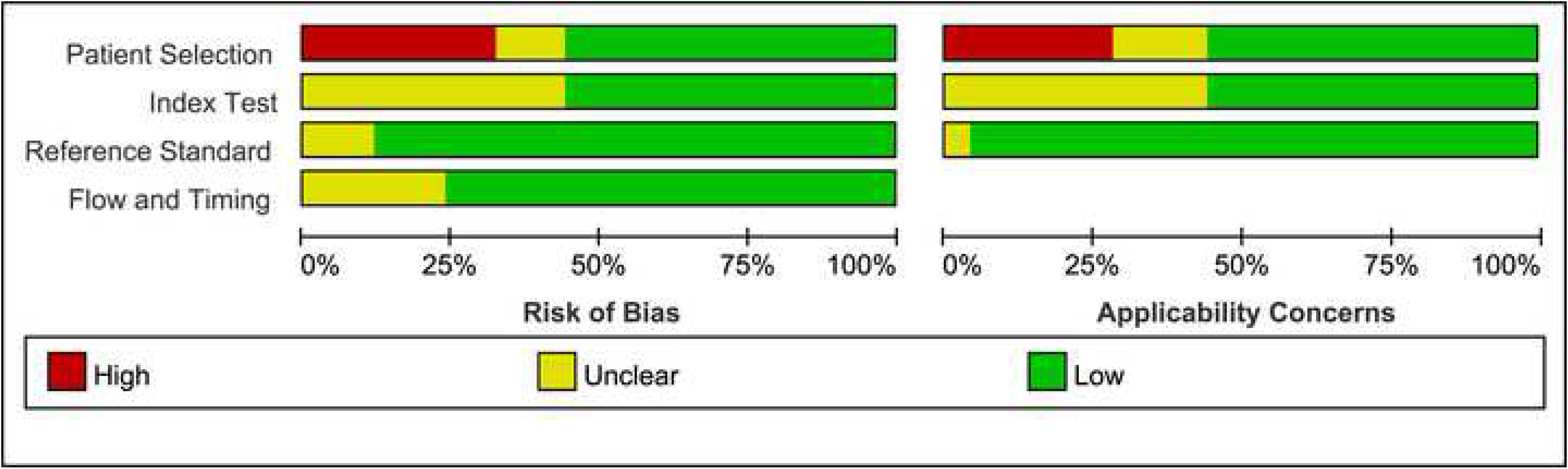
Quality Assessment of Diagnostic Accuracy Studies (QUADAS)-2 graphs of CT in diagnosing COVID-19.

We evaluated the publication bias of 11 studies, which detailed the diagnostic performance of CT (true positive, false positive, true negative, and false negative). As a result, the funnel plot appeared to be symmetrical, with a P-value of 0.87, indicating a low risk of publication bias (Figure 3).

**Figure 3:**
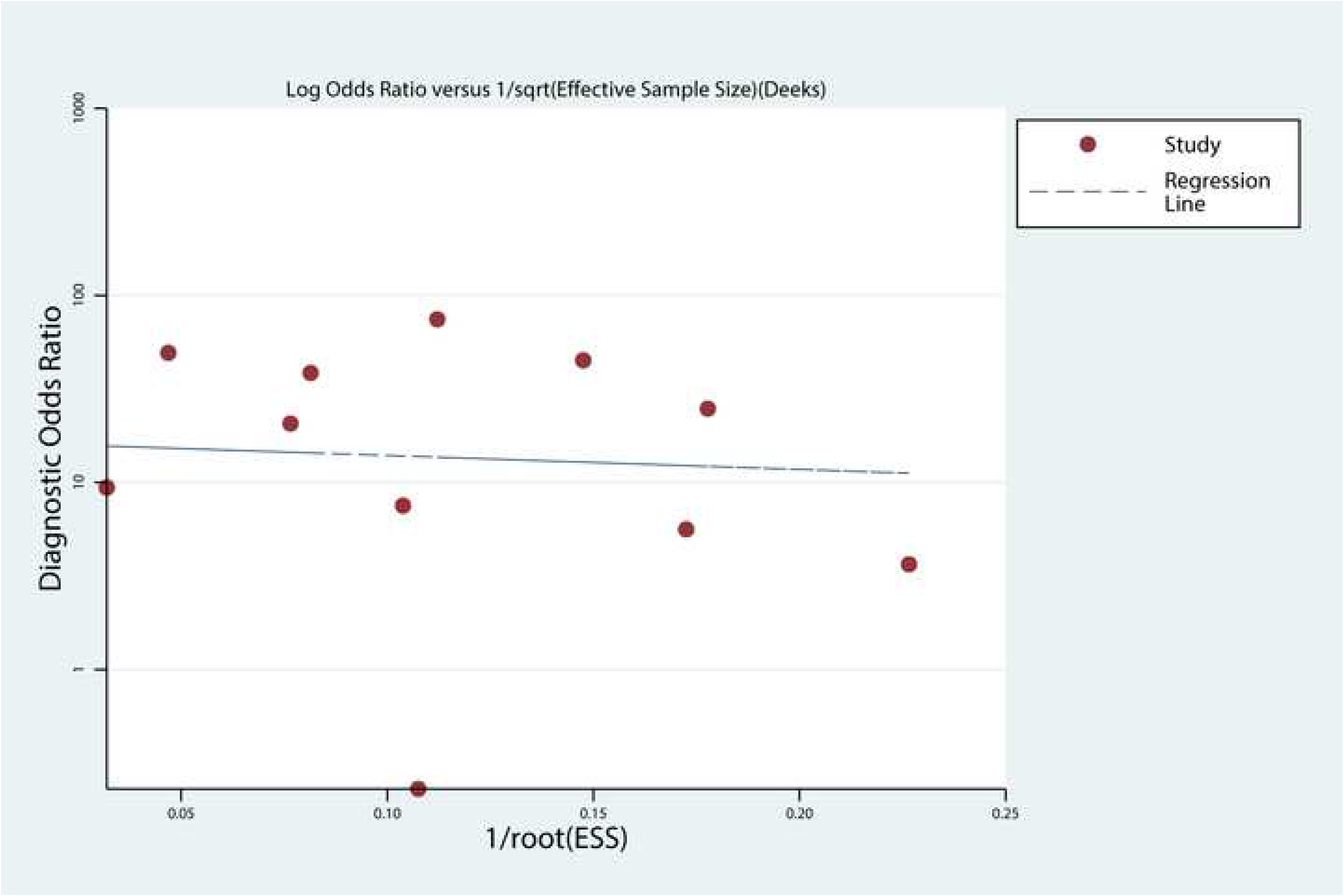
Deeks’ funnel plot of 11 included studies (p = 0.87 in Deeks’ asymmetry test).

### 3.3 Meta-analysis results

#### 3.31 Demographical characteristics and Clinical features

The mean or median age of patients diagnosed with COVID-19 ranged from 41 to 58, and the proportion of males was 57% (95%CI 62-72%). A total of 81% (95%CI 72-89%) of patients had an epidemiological history. The incidence of fever was 79% (95%CI 66-90%), and that of cough was 58% (95%CI 48-68%).

#### 3.32 Pooled Diagnostic Performance

For these 25 studies, the diagnostic sensitivity and specificity of CT for COVID-19 ranged from 69% to 100% and from 0% to 96%, with pooled estimates of 93% (95% CI, 89-96%) and 44% (95% CI, 27-62%) (Figure 4), respectively. The pooled positive predictive value and negative predictive value of CT for COVID-19 were 0.55 (95% CI, 0.41-0.68) and 0.97 (95% CI, 0.91-1.00), respectively. The Q-test P<0.01 and I^2^>50% indicated heterogeneity of sensitivity and specificity between studies. We plotted the SROC curve to display the diagnosis results graphically and the area under the SROC curve was 0.94 (95% CI, 0.91-0.96). The SROC curve did not suggest that there is a threshold effect (Figure 5).

**Figure 4:**
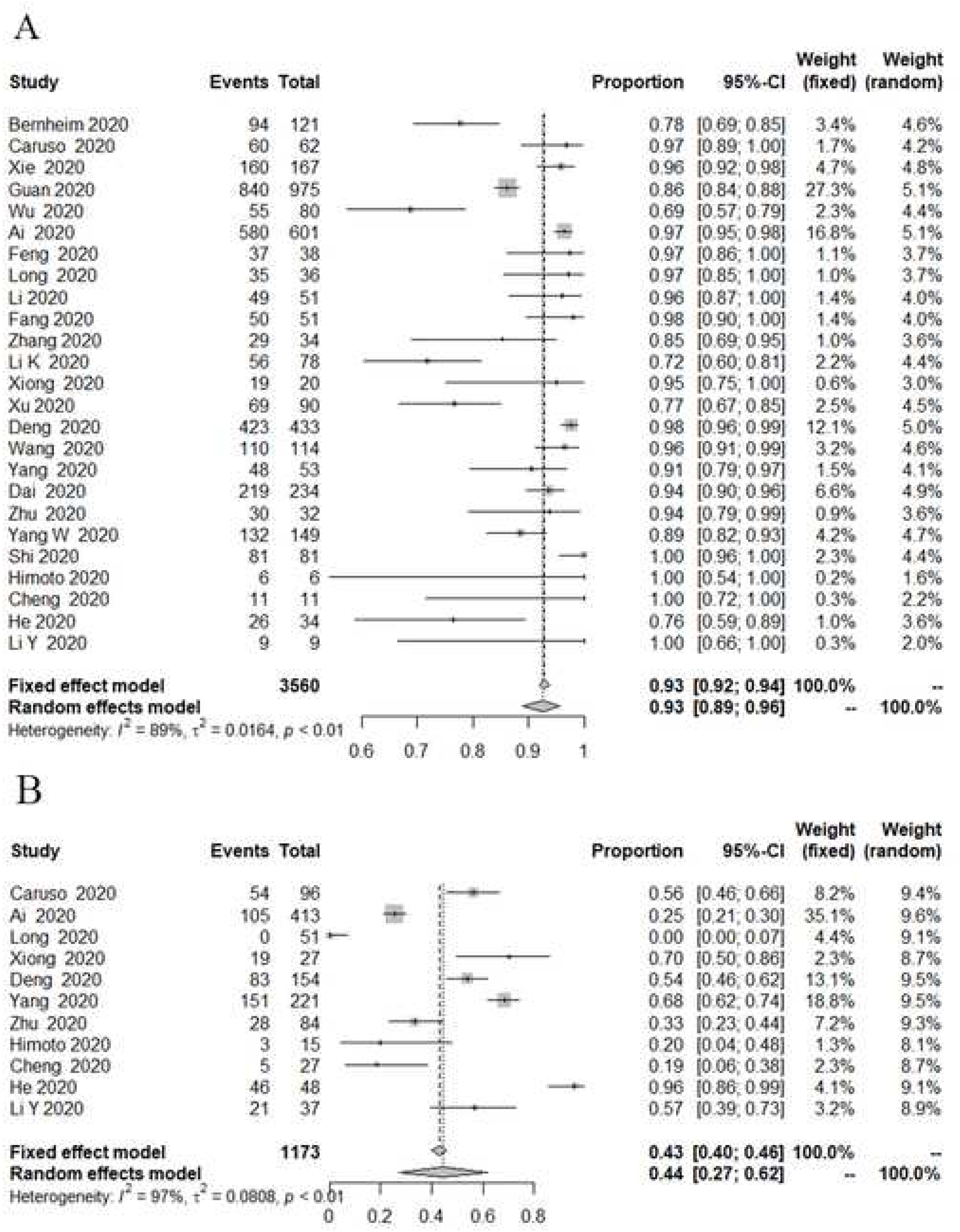
Forest plots of the diagnostic performance of chest CT. A: A plot of 25 individual studies and pooled sensitivity of chest CT in patients with suspicious COVID-19. B: A plot of 11 individual studies and pooled specificity of chest CT in patients with suspicious COVID-19.

**Figure 5:**
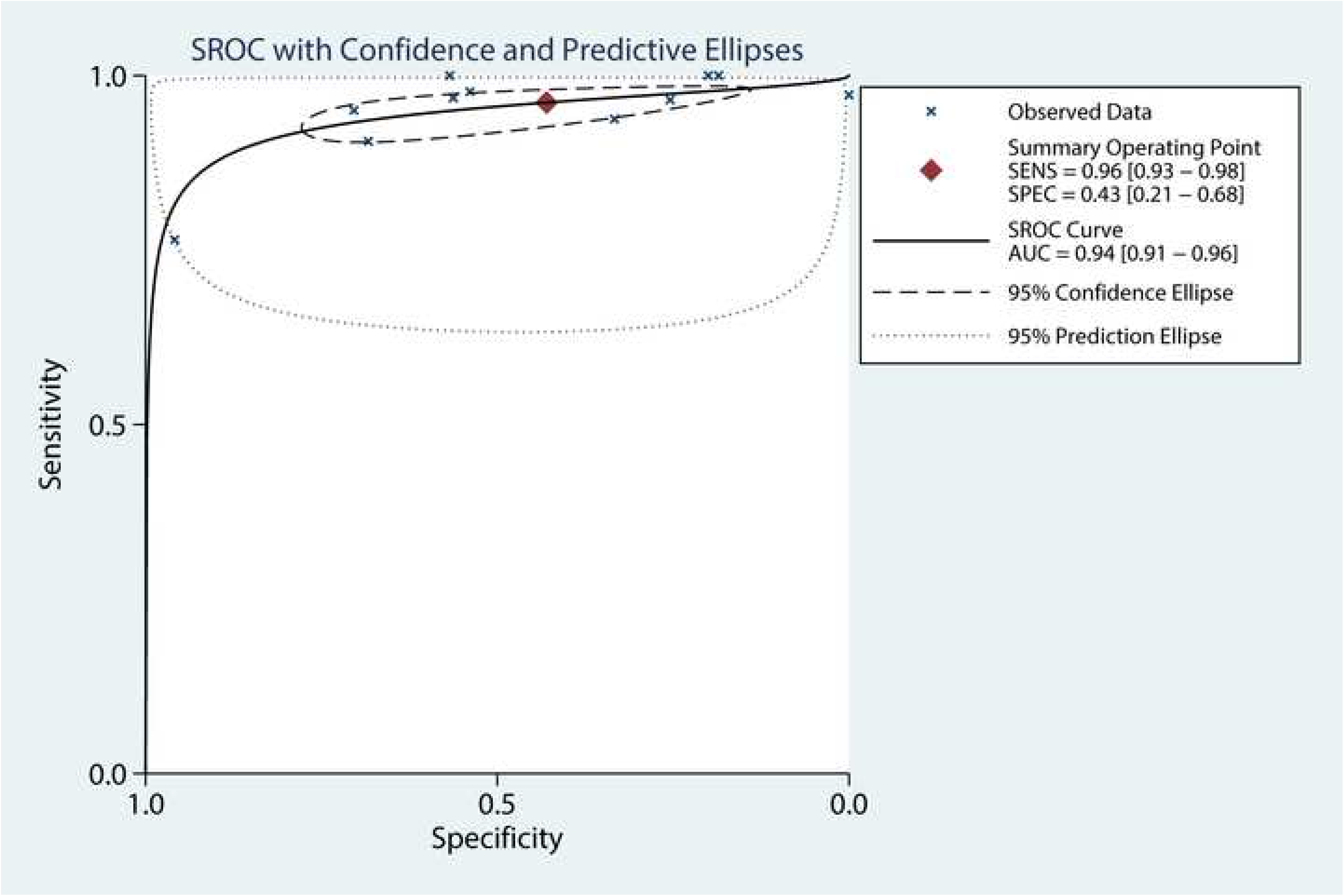
Summary receiver operating curves (SROC) for the diagnostic performance of CT.

For the 10 studies with repeated PCR assay, the pooled sensitivity of the initial RT-PCR test in diagnosis of COVID-19 was 76% (95% CI: 59-89%; I^2^=96%). Besides, 5 studies conducted three or more PCR tests for patients with detailed records of each positive result. For suspected cases with initially negative RT-PCR test, the missed diagnosis rate of COVID-19 when only took second-round RT-PCR examination was 26% (95% CI: 14-39%; I^2^=45%) (Figure 6).

**Figure 6:**
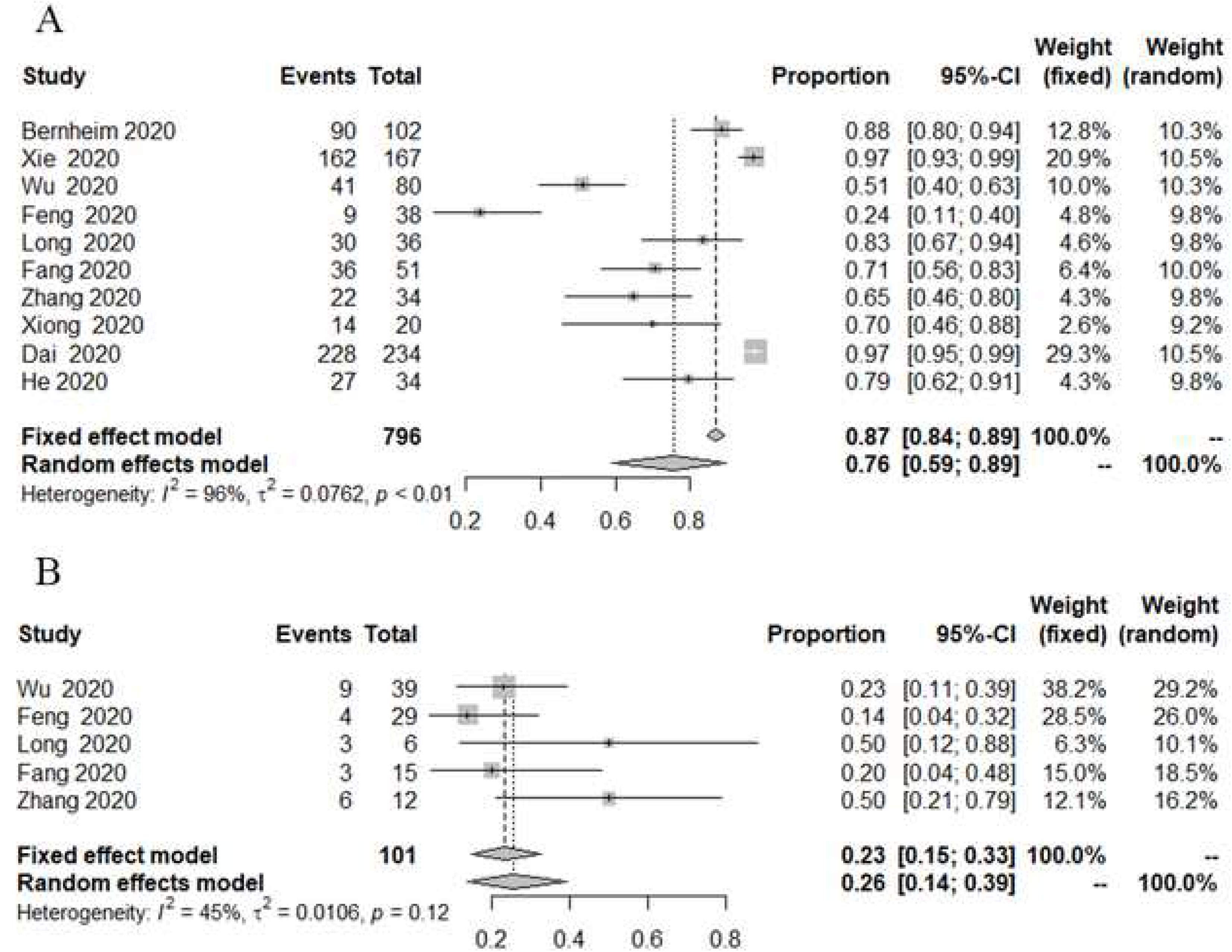
Forest plots of the diagnostic performance of chest CT. A: A plot of 10 individual studies and pooled sensitivity of the initial RT-PCR assay in patients with suspicious COVID-19. B: A plot of 5 individual studies and pooled rate of missed diagnosis after the second-round RT-PCR examinations.

#### 3.33 Imaging features of Chest CT

GGO was a key sign of CT imaging, and its incidence was 58% (95% CI: 49-70%) in patients with SARS-CoV-2 infection. Other common signs included air bronchogram 51% (95% CI: 31-70%), interlobular septal thickening 32% (95% CI: 17-49%), airways abnormalities 31% (95% CI: 7-61%), linear opacities 29% (95% CI: 3-66%), consolidation 20% (95%CI: 7-37%), and crazy-paving pattern 17% (95 CI: 4-36%). We found that nodules, pleural effusion and lymphadenopathy are less frequent findings in this meta-analysis, and their incidences were 5% (95% CI: 1-12%), 4% (95% CI: 2-6%) and 4% (95% CI: 0-10%), respectively.

Pneumonia lesions were inclined to distribute in peripheral 64% (95% CI: 49-78%) and bilateral 69% (95% CI: 58-79%) lung lobes. The incidence of two or more lobes affected and a simultaneous involvement of all five lobes was 61% (95% CI: 48-74%) and 48% (95% CI: 30-66%), respectively. For each lung lobe, the right lower lobe and the left lower lobe had the highest frequency of involvement, with an incidence of 51% (95% CI: 35-67%) and 49% (95% CI: 34-64%), respectively.

### 3.4 meta-regression analyses

The results of heterogeneity exploration were presented in Table 4. Among several covariates assessed for the sensitivity of chest CT, only the study region significantly affected the heterogeneity (p <0.05). Moreover, the sample size and time interval between reference standard and CT did not show an effect on CT sensitivity (p = 0.68-0.84). For the diagnostic sensitivity of CT, none of the three variables showed significant effect on heterogeneity (p = 0.07-0.81). Both regions and sample sizes did not affect the premier sensitivity of RT-PCR (p = 0.09-0.39).

**Table 4:** Results of meta-regression analysis. 95% CI = 95% confidence interval.

| Variable | No.<br>of studies | Pooled Sensitivity<br>of CT (95% CI) | P value | No.<br>of studies | Pooled Specificity<br>of CT (95% CI) | P value | No.<br>of studies | Pooled Sensitivity of<br>initial RT-PCR (95% CI) | P value |
| --- | --- | --- | --- | --- | --- | --- | --- | --- | --- |
| Hubei/China |  |  | 0.03 |  |  | 0.07 |  |  | 0.39 |
| Yes | 8 | 0.96 (0.92; 0.99) |  | 3 | 0.22 (0.02; 0.54) |  | 2 | 0.54 (0.04; 0.99) |  |
| No | 17 | 0.90 (0.84; 0.95 ) |  | 8 | 0.54 (0.37; 0.71) |  | 8 | 0.81 (0.65; 0.92) |  |
| Sample size |  |  | 0.84 |  |  | 0.81 |  |  | 0.09 |
| ≥100 | 12 | 0.92 (0.87; 0.95) |  | 5 | 0.47 (0.29; 0.66) |  | 4 | 0.87 (0.68; 0.99) |  |
| <100 | 13 | 0.94 (0.87; 0.99) |  | 6 | 0.41 (0.06; 0.82) |  | 6 | 0.66 (0.47; 0.82) |  |
| CT-Reference interval |  |  | 0.68 |  |  | 0.43 | - | - | - |
| Mentioned (≤2 weeks) | 8 | 0.93 (0.87; 0.98) |  | 4 | 0.35 (0.06; 0.73) |  | - | - | - |
| Not mentioned | 17 | 0.92 (0.88; 0.96) |  | 7 | 0.51 (0.39; 0.63) |  | - | - | - |

## 4. Discussion

In this study, the meta-analysis was used to quantify the diagnostic performance of CT for COVID-19. We found that the comprehensive diagnostic performance of CT for COVID-19 was good. Specifically, the pooled sensitivity and specificity were 93% (95% CI, 89-96%) and 44% (95% CI, 27-62%), respectively, with an AUC of 0.94 (95% CI,0.91-0.96). However, the diagnostic specificity of CT for patients suspected of being infected with SARS-CoV-2 was low. Our results showed that the pooled sensitivity of the initial RT-PCR assay was 76% (95% CI: 59-89%; I^2^=96%). It is worth noting that even after taking second-round PCR tests, about 26% (95% CI: 14-39%) of patients who have been infected with coronavirus will still be missed. The positive findings of chest CT images suggest acute alveolitis, which is a similar common manifestation of various viral pneumonia, partial bacterial pneumonia and lung damage caused by immune diseases, but not a specific sign of COVID-19 [38]. COVID-19 is an infectious disease, which still needs to be confirmed by etiological testing. Because imaging features of CT might be similar in different diseases, it could not replace the etiological examination in the diagnosis of COVID-19.

Although the specificity of CT was not high, our results in this meta-analysis indicated that CT was indispensable in screening patient with 2019 novel coronavirus when considering the following factors: (1) given the current global outbreak of COVID-19, RT-PCR testing is time-consuming and the kits in some areas are insufficient, which will lead to the gathering of people in emergency departments and the flow of sources of infection. CT examination has the advantages of rapid, objective, and reproducible; (2) there may be false-negative cases in RT-PCR assay, so using the RT-PCR for screening alone may result in a certain number of missed diagnoses. The high sensitivity of CT can compensate for the deficiency of RT-PCR screening; (3) PCR testing requires a complex sample collection process. Some patients have more virus in the nasopharynx and some in the oropharynx, while a few patients can only find the virus in bronchoalveolar lavage fluid, so it is prone to sampling errors; (4) different CT image manifestations can be used to assess the clinical course and severity of the disease[39]. Of course, there are some shortcomings in CT inspection, including poor air permeability in the operating room and cumbersome disinfection. These may lead to potential cross-infection. Therefore, we can consider adopting time-sequence, low-to-high risk order, and strict disinfection to reduce the risk.

Ground-glass opacities were the most typical manifestation in this study. Most of the lesions showed peripheral distribution with or without interlobular septal thickening. The mechanism of these changes may be the fluid exudation in the alveolar cavity caused by telangiectasia of the alveolar septum and interstitial edema of the interlobular septum [40]. In addition, our study showed that the incidence of air bronchogram is high, and consolidation is relatively rare. However, the main feature of pulmonary manifestations in SARS was large pulmonary consolidation, which was often accompanied by obvious air bronchogram [41]. Liu et al analyzed the CT findings of 73 patients with SARS-CoV-2 infection and found that pulmonary consolidation mainly occurred in severe and critically ill patients [40]. This meta-analysis suggested that about 4% of patients had pleural effusion, which might be a sign of a poor prognosis [42]. COVID-19 also needs to be distinguished from MERS (Middle East respiratory syndrome). Both of them showed multiple ground glass shadow lesions under the bilateral pleura [43].

There was considerable heterogeneity between the literature included in our study. According to the results of meta-regression analysis, in high-prevalence regions, the diagnostic sensitivity of CT for COVID-19 was significantly higher than that in low-prevalence regions. It meant that CT could be used as an effective screening tool for COVID-19 outbreak areas. However, we have not yet been able to explore the factors that affect the specificity of CT and the sensitivity of the initial RT-PCR test. More studies are needed to confirm the involved factors, such as the internal diagnostic thresholds of chest CT imaging, the turn-around time of sample in transportation, and the type of kits.

Our review had several limitations. Only two documents came from non-Chinese regions, and the detailed information of CT in different regions was lacking. Information about the diagnostic specificity of CT could not be obtained from most studies. Therefore, we only included 11 articles with 2 × 2 tables to draw the SROC curve. Some studies did not mention the time of CT and RT-PCR examination. When exploring heterogeneity, the literature with the interval between CT and PCR examinations less than two weeks was divided into one group, and the rest constituted another group. It was best to obtain the specific time interval information to fully understand the factors influencing diagnostic sensitivity of CT. In addition, the study did not obtain sufficient CT image data to explain the relationship between imaging manifestations and duration of Infection.

In conclusion, CT was highly sensitive in the diagnosis of COVID-19. There were still several cases of missed diagnosis after multiple RT-PCR examinations. In high-prevalence areas, CT can be recommended as an auxiliary screening method for RT-PCR. In the future, large samples and high-quality prospective studies can make up for the deficiency of the current small sample size and further verify the results.

## Data Availability

Statement regarding the availability of all data was not required for this study because the article type is a meta-analysis.

## Acknowledgments

The authors state that this work has not received any funding.

## Declaration of Competing Interest

The authors in this study have no conflict of interest.

**Abbreviations**
COVID-19: 2019 novel coronavirus disease
RT-PCR: reverse transcription polymerase chain reaction
SROC: summary receiver operating characteristic
GGO: ground glass opacities
TP: true positive
FP: false positive
FN: false negative
TN: true negative
AUC: area under the curves

## Reference

1. Huang C, Wang Y, Li X, Ren L, Zhao J, Hu Y, et al. (2020) Clinical features of patients infected with 2019 novel coronavirus in Wuhan, China. Lancet. 395(10223):497–506.

2. Lu R, Zhao X, Li J, Niu P, Yang B, Wu H, et al. (2020) Genomic characterisation and epidemiology of 2019 novel coronavirus: implications for virus origins and receptor binding. Lancet. 395(10224):565–574.

3. Wu Z, McGoogan JM. (2020) Characteristics of and Important Lessons From the Coronavirus Disease 2019 (COVID-19) Outbreak in China: Summary of a Report of 72 314 Cases From the Chinese Center for Disease Control and Prevention. JAMA 10.1001/jama.2020.2648

4. World Health Organization (2020) main website. Available via https://www.who.int. Accessed 23 April 2020.

5. Al-Tawfiq J A, Memish Z A. (2020) Diagnosis of SARS-CoV-2 Infection based on CT scan vs. RT-PCR: Reflecting on Experience from MERS-CoV. The Journal of hospital infection 10.1016/j.jhin.2020.03.001

6. Jin YH, Cai L, Cheng ZS, Cheng H, Deng T, Fan YP, et al. (2020) A rapid advice guideline for the diagnosis and treatment of 2019 novel coronavirus (2019-nCoV) infected pneumonia (standard version). Mil Med Res 7(1):4.

7. Xie X, Zhon Z, Zhao W, Zheng C, Wang F, Liu J. (2020) Chest CT for Typical 2019-nCoV Pneumonia: Relationship to Negative RT-PCR Testing. Radiology 10.1148/radiol.2020200343:200343.

8. Huang P, Liu T, Huang L, Liu H, Lei M, Xu W, et al. (2020) Use of Chest CT in Combination with Negative RT-PCR Assay for the 2019 Novel Coronavirus but High Clinical Suspicion. Radiology.295(1):22–23.

9. Caruso D, Zerunian M, Polici M, Pucciarelli F, Polidori T, Rucci C, et al. (2020) Chest CT Features of COVID-19 in Rome, Italy. Radiology 10.1148/radiol. 2020 201237: 201237.

10. Fang Y, Zhang H, Xie J, Lin M, Ying L, Pang P, et al. (2020) Sensitivity of Chest CT for COVID-19: Comparison to RT-PCR. Radiology 10.1148/radiol. 2020 200432: 200432.

11. Bernheim A, Mei X, Huang M, Yang Y, Fayad ZA, Zhang N, et al. (2020) Chest CT Findings in Coronavirus Disease-19 (COVID-19): Relationship to Duration of Infection. Radiology 10.1148/radiol.2020200463:200463.

12. Hope MD, Raptis CA, Shah A, Hammer MM, Henry TS (2020) A role for CT in COVID-19? What data really tell us so far. Lancet. 395(10231):1189–1190.

13. PROSPERO (2020) CRD42020179689. Available via https://www.crd.york.ac.uk/PROSPERO/ Accessed 16 April 2020.

14. Whiting PF, Rutjes AW, Westwood ME, Mallett S, Deeks JJ, Reitsma JB et al. (2011) QUADAS-2: a revised tool for the quality assessment of diagnostic accuracy studies. Ann. Intern. Med. 155 (8):529–536.

15. Luo ML, Tan HZ, Zhou Q, Wang SY, Cai C, Guo YW, et al. (2013) Realizing the Meta-Analysis of Single Rate in R Software. The Journal of Evidence-Based Medicine.13(3).

16. Lee J, Kim KW, Choi SH, Huh J, Huh J (2015) Systematic Review and Meta-Analysis of Studies Evaluating Diagnostic Test Accuracy: A Practical Review for Clinical Researchers-Part II. Statistical Methods of Meta-Analysis. Korean J Radiol. 16(6):1188.

17. Deng ZQ, Zhang CX, Li YR, Xu HB, Gang YD, Wang HL, et al. (2020) Value of chest CT screening in the early COVID-19 outbreak Chin J Radiol. 54.

18. Xiong Z, Fu L, Zhou H, Liu JK, Wang AM, Huang Y, et al. (2020) Construction and evaluation of a novel diagnosis process for 2019-Corona Virus Disease. Zhonghua yi xue za zhi. 100:E019.

19. Cheng Z, Lu Y, Cao Q, Qin L, Pan Z, Yan F, et al. (2020) Clinical Features and Chest CT Manifestations of Coronavirus Disease 2019 (COVID-19) in a Single-Center Study in Shanghai, China. AJR. American journal of roentgenology 10.2214/AJR.20.22959:1–6.

20. Li Y, Xia L (2020) Coronavirus Disease 2019 (COVID-19): Role of Chest CT in Diagnosis and Management. AJR. American journal of roentgenology 10.2214/AJR.20.22954:1–7.

21. Himoto Y, Sakata A, Kirita M, Hiroi T, Kobayashi KI, Kubo K, et al. (2020) Diagnostic performance of chest CT to differentiate COVID-19 pneumonia in non-high-epidemic area in Japan. Jpn J Radiol 10.1007/s11604–020-00958-w.

22. Feng Y, Yuan LF, Zheng CX, Liu WS, Wang YJ, Ren CY, et al. (2020) Practical application of CT and nucleic acid detection in the diagnosis of COVID-19. Guangdong Medical Journal 10. 13820/j. cnki. Gdyx. 20200302.

23. Yang XL, Wang ZH, Liu X, Wu SS, Wu XP, Wen GL, et al. (2020) Screening for 274 suspected cases of novel coronavirus pneumonia 38.

24. Xu X, Yu C, Qu J, Zhang L, Jiang S, Huang D, et al. (2020) Imaging and clinical features of patients with 2019 novel coronavirus SARS-CoV-2. European Journal of Nuclear Medicine and Molecular Imaging 10.1007/s00259–020-04735-9

25. Guan WJ, Ni ZY, Hu Y, Liang W.H, Ou CQ, He JX, et al. (2020) Clinical Characteristics of Coronavirus Disease 2019 in China. The New England journal of medicine 10.1056/NEJMoa2002032.

26. Zhu W, Xie K, Lu H, Xu L, Zhou S, Fang S (2020) Initial clinical features of suspected coronavirus disease 2019 in two emergency departments outside of Hubei, China. J Med Virol 10.1002/jmv.25763.

27. Yang W, Cao Q, Qin L, Wang X, Cheng Z, Pan A, et al. (2020) Clinical characteristics and imaging manifestations of the 2019 novel coronavirus disease (COVID-19):A multi-center study in Wenzhou city, Zhejiang, China. Journal of Infection 10.1016/j.jinf.2020.02.016.

28. Ai T, Yang Z, Hou H, Zhan C, Chen C, Lv W, et al. (2020) Correlation of Chest CT and RT-PCR Testing in Coronavirus Disease 2019 (COVID-19) in China: A Report of 1014 Cases. Radiology 10.1148/radiol.2020200642:200642.

29. Zhang SX, Li J, Zhou P, Na JR, Liu BF, Zheng XW, et al. (2020) The analysis of clinical characteristics of 34 novel coronavirus pneumonia cases in Ningxia Hui autonomous region. 43.

30. Wang K, Kang S, Tian R, Zhang X, Wang Y (2020) Imaging manifestations and diagnostic value of chest CT of coronavirus disease 2019 (COVID-19) in the Xiaogan area. Clinical Radiology 10.1016/j.crad.2020.03.004

31. Li K, Fang Y, Li W, Pan C, Qin P, Zhong Y, et al. (2020) CT image visual quantitative evaluation and clinical classification of coronavirus disease (COVID-19). European radiology 10.1007/s00330–020-06817-6

32. Wu J, Liu J, Zhao X, Liu C, Wang W, Wang D, et al. (2020) Clinical Characteristics of Imported Cases of COVID-19 in Jiangsu Province: A Multicenter Descriptive Study. Clinical infectious diseases: an official publication of the Infectious Diseases Society of America 10.1093/cid/ciaa199

33. Shi H, Han X, Jiang N, Cao Y, Alwalid O, Gu J, et al. (2020) Radiological findings from 81 patients with COVID-19 pneumonia in Wuhan, China: a descriptive study. The Lancet Infectious Diseases.20(4):425–434.

34. Dai H, Zhang X, Xia J, Zhang T, Shang Y, Huang R, et al. (2020) High-resolution Chest CT Features and Clinical Characteristics of Patients Infected with COVID-19 in Jiangsu, China. 10.1016/j.ijid.2020.04.003

35. Long C, Xu H, Shen Q, Zhang X, Fan B, Wang C, et al. (2020) Diagnosis of the Coronavirus disease (COVID-19): rRT-PCR or CT? European Journal of Radiology. 126.

36. He JL, Luo L, Luo ZD, Lyu JX, Ng MY, Shen XP, et al. (2020) Diagnostic performance between CT and initial real-time RT-PCR for clinically suspected 2019 coronavirus disease (COVID-19) patients outside Wuhan, China. 168:105980.

37. Li Y, Xu SY, Du TK, Xu J, Li Y, Yu XZ, et al. (2020) Clinical features of 2019 novel coronavirus infection patients and a feasible screening procedure Chin J Emerg Med 29 E007-E007.

38. Jin YH, Chen B, Zhang JF, Dai Q, Yan K, Ye HH, et al. (2020) The value of chest CT Imaging in the Prevention and Control of COVID-19 epidemic situation. Journal of Modern practical Medicine.32(2)

39. Wu J, Wu X, Zeng W, Guo D, Fang Z, Chen L, et al. (2020) Chest CT Findings in Patients with Corona Virus Disease 2019 and its Relationship with Clinical Features. Investigative radiology 10.1097/RLI.0000000000000670

40. Liu KC, Xu P, Lv WF, Qiu XH, Yao JL, Gu JF, et al. (2020) CT manifestations of coronavirus disease-2019: A retrospective analysis of 73 cases by disease severity. Eur J Radiol.126:108941.

41. Wang JT, Sheng WH, Fang CT, Chen YC, Wang JL, Yu CJ, et al. (2004) Clinical manifestations, laboratory findings, and treatment outcomes of SARS patients. Emerg Infect Dis.10:818–824.

42. Das KM, Lee EY, Al Jawder SE, Enani MA, Singh R, Skakni L, et al. (2015) Acute Middle East Respiratory Syndrome Coronavirus: Temporal Lung Changes Observed on the Chest Radiographs of 55 Patients. 205(3): W267–274.

43. Ajlan AM, Ahyad RA, Jamjoom LG, Alharthy A, Madani TA (2014) Middle East respiratory syndrome coronavirus (MERS-CoV) infection: chest CT findings. American journal of roentgenology 203(4):782–787.

